# Increased household transmission and immune escape of the SARS-CoV-2 Omicron variant compared to the Delta variant: evidence from Norwegian contact tracing and vaccination data

**DOI:** 10.1101/2022.02.07.22270437

**Authors:** Neda Jalali, Hilde K. Brustad, Arnoldo Frigessi, Emily MacDonald, Hinta Meijerink, Siri Feruglio, Karin Nygård, Gunnar Isaksson Rø, Elisabeth H. Madslien, Birgitte Freiesleben De Blasio

## Abstract

Understanding the rapid epidemic growth of the novel SARS-CoV-2 Omicron variant is critical for public health management. We compared the secondary attack rate (SAR) of the Omicron and Delta variants in households using Norwegian contact tracing data from December 2021 to January 2022. Omicron SAR was higher (51%) than Delta (36%), with a relative risk (RR) of 1.41 (95% CI 1.27-1.56). We observed increased susceptibility to Omicron infection in household contacts compared to Delta independent of vaccination status; however, considering booster vaccinated contacts, the mean SAR was lower for both variants. We found increased Omicron transmissibility in all vaccination groups of primary cases, except partially vaccinated, compared to Delta. In particular, Omicron SAR for boosted primary cases was high, 46% vs 11 % for Delta (RR 4.34; 95% CI 1.52-25.16). In conclusion, booster doses decrease the infection risk of Delta and Omicron but have limited effect in preventing Omicron transmission.

## INTRODUCTION

By the end of 2021, rapid global spread of the novel SARS-CoV-2 Omicron variant of concern (VoC) (Pangolin designation B.1.1.529 BA.1) caused major concern and urgent need for knowledge about its transmissibility, severity of disease and ability to escape vaccine immunity [1, 2] In Norway, the first SARS-CoV-2 Omicron case was reported on 30 November at a time when the Delta variant was dominating. At the same time, a large Omicron outbreak was detected after a Christmas party, causing an attack rate of 74% among participants, of which most (98%) were fully vaccinated [3]. Over the next 4-6 weeks, Omicron rapidly took over for the Delta variant in Norway and by week 2 in 2022 the Omicron variant was detected in > 90% of the weekly national samples screened or sequenced for virus variants [4]. Early studies report that Omicron might have higher transmissibility than Delta [5-7], although infection seems to cause less severe disease and lower risk of hospitalization [1, 8]. The Omicron variant’s ability to escape vaccine immunity [9-11] is likely an important contributor to the current rapid spread of the disease [7]. A highly immune evasive VoC could potentially challenge global control strategies. Thus, timely and relevant knowledge about transmissibility and risk of infection concerning vaccination status of the population is of particular importance to guide health authorities.

Here, we use contact tracing data collected by Norwegian municipalities to estimate and compare household secondary attack rate (SAR) for the Omicron and Delta variants at a time when both variants were circulating throughout the country.

## METHODS

### Study design, study population and data sources

We conducted a registry-based cohort study using data from the Norwegian COVID-19 pandemic preparedness register, Beredt C19 [12]. Beredt C19 receives individual-level information from Norwegian health registries which can be linked using unique personal identification numbers. The purpose of the preparedness register is to enable rapid knowledge generation on the spread of COVID-19 to support national authorities in crisis management and preparedness planning. Beredt C19 collects information from various national registries, including information on Norwegian residents who have tested positive for SARS-CoV-2, date of testing, variant detection, vaccination record, and demographics. Furthermore, the register receives digital contact tracing data on a voluntary basis from Norwegian municipalities, enabling the linkage of index cases to their traced contacts. Detailed information on the specific Beredt C19 data sources that were used in this study is shown in Supplementary S1.

The study population were households registered in the municipal contact tracing system of 64 municipalities within the study period. We defined a primary case as the first person in a household (according to the testing date) to test positive for either the SARS-CoV-2 Omicron or Delta variant during the study period. Virus variant information was based on either PCR variant screening, whole genome sequencing, or both. Household contacts of the primary cases were identified by matching household identification numbers of the primary case and contacts. We limited our study to households of sizes between 2 – 6 individuals to exclude multi-generation households, care facilities and institutions. Furthermore, six primary cases and 36 contacts were excluded from the analysis due to previously reported SARS-CoV-2 infection. We defined a household secondary case as any individual registered as a close contact of the primary case, living in the same household and who tested positive for SARS-CoV-2 ≤ 10 days after the test date of the primary case. Vaccination status of the cases and contacts were separated into the following categories: i) unvaccinated, ii) partially vaccinated, iii) fully vaccinated and iv) booster vaccinated. Further information on definitions used in the study is described in Supplementary S2. We limited data collection to the period from 14 December 2021 to 23 January 2022 to avoid bias due to differences in the TISK regime for Omicron and Delta. We included only primary cases who tested positive until 13 January 2022, to ensure the maximum of ten days contact tracing is satisfied for all the cases.

### Statistical Analysis

We used binomial regression with a log link to estimate the secondary attack rate (SAR) within households, comparing Delta with Omicron, assuming test activity and case finding did not vary by variant [13]. The binomial regression model was stratified for different covariates, including vaccination status, age group, and gender, of the contacts and primary cases to find the relative risk (RR) between them. Vaccine effectiveness (VE) against infection among household contacts 16 years and above was calculated using the following equation: VE= 1-(SAR_vaccinated contacts_/SAR_unvaccinated contacts_). Contacts aged 0-15 years were excluded from the VE-calculations because in Norway children 12-15 years were only eligible for one vaccine dose, while young children 0-11 years were not offered vaccination at the time. Significance level (α) was set at 5%. Statistical analyses were performed with Rstudio 1.3.1056.

## RESULTS

In total, 1122 primary cases with confirmed Delta (41%) or Omicron (59%) and 2169 household contacts (60% for Omicron primary cases, and 40% for Delta primary cases) representing 57/356 municipalities and 8/11 counties were registered in the contact tracing system and included in the final dataset. Characteristics of the cases and contacts are presented in Supplementary S3.

The main findings are presented in Table 1A, B, C. The overall SAR of households with Omicron was estimated at 51% (CI_95_: 48-54) compared to 36% (CI_95_: 33-40) with Delta giving a significantly higher risk of infection in households with Omicron relative to Delta (Table 1A). In both age groups, a primary case infected with Omicron has significantly higher risk of COVID-19 transmission versus a primary case infected by Delta. The risk of infection in all vaccination groups of contacts was significantly higher in households with Omicron relative to Delta (Table 1A). Generally, the SAR in households with booster vaccinated primary cases and contacts was lower than in households with unvaccinated primary cases and contacts. Primary cases who were booster vaccinated were found to have a considerably higher risk (RR: 4.34; CI_95_: 1.52-25.16) of transmitting SARS-CoV-2 to their household contacts with Omicron compared to Delta. A similar trend was observed when the primary case was unvaccinated or fully vaccinated, although the relative risks were lower (Unvaccinated: RR: 1.51; CI_95_: 1.30-1.77, Fully vaccinated: RR: 1.44; CI_95_: 1.24-1.70).

**Table 1:** Secondary attack rates (SAR) and relative risks (RR) of Omicron versus Delta stratified by contacts’ age, primary cases’ age, vaccination status of contacts, primary cases, gender, and time since last vaccination in fully vaccinated contacts in Norway, 14 December 2021 to 23 January 2022.

| **Table** | **Description** | **Omicron** |  | **Delta** |  |
| --- | --- | --- | --- | --- | --- |
| **1-A** | **SAR, between variant RR** | **SAR (95% CI)** | **RR (95% CI)** | **SAR (95% CI)** | **RR** |
|  | Overall | 0.51 (0.48 - 0.54) | **1.41 (1.27 - 1.56)** | 0.36 (0.33 - 0.40) | Ref |
|  | Age of contacts |  |  |  |  |
|  | <16 years | 0.58 (0.53 - 0.63) | **1.38 (1.18 - 1.64)** | 0.42 (0.36 - 0.48) | Ref |
|  | 16+ years | 0.49 (0.45 - 0.51) | **1.44 (1.27 - 1.65)** | 0.33 (0.30 - 0.37) | Ref |
|  | Age of primary cases |  |  |  |  |
|  | <16 years | 0.53 (0.48 - 0.58) | **1.53 (1.29 - 1.82)** | 0.35 (0.30 - 0.40) | Ref |
|  | 16+ years | 0.50 (0.47 - 0.53) | **1.35 (1.19 - 1.54)** | 0.37 (0.33 - 0.41) | Ref |
|  | Vacc. status, contacts |  |  |  |  |
|  | Unvaccinated | 0.59 (0.53 - 0.64) | **1.27 (1.09 - 1.48)** | 0.47 (0.41 - 0.52) | Ref |
|  | Partial | 0.57 (0.48 - 0.66) | **1.69 (1.18 - 2.55)** | 0.34 (0.23 - 0.46) | Ref |
|  | Full | 0.50 (0.46 - 0.54) | **1.55 (1.33 - 1.82)** | 0.32 (0.28 - 0.37) | Ref |
|  | Boosted | 0.38 (0.31 - 0.44) | **1.91 (1.19 - 3.40)** | 0.20 (0.11 - 0.30) | Ref |
|  | Vacc. status, primary cases |  |  |  |  |
|  | Unvaccinated | 0.57 (0.51 - 0.62) | **1.51 (1.30 - 1.77)** | 0.38 (0.33 - 0.42) | Ref |
|  | Partial | 0.42 (0.34 - 0.49) | 1.05 (0.76 - 1.52) | 0.39 (0.29 - 0.51) | Ref |
|  | Full | 0.51 (0.47 - 0.55) | **1.44 (1.24 - 1.70)** | 0.35 (0.31 - 0.40) | Ref |
|  | Boosted | 0.46 (0.36 - 0.55) | **4.34 (1.52 - 25.16)** | 0.11 (0.02 - 0.29) | Ref |
| **1-B** | **VE by contact vacc. status (16+)** | **Omicron VE % (95% CI)** |  | **Delta VE % (95% CI)** |  |
|  | Unvaccinated | Ref |  | Ref |  |
|  | Partial | 22 (0 - 46) |  | 31 (0 - 72) |  |
|  | Full | 27 (6 - 49) |  | 42 (23 - 55) |  |
|  | Boosted | 45 (26 - 57) |  | 65 (42 - 80) |  |
| **1-C** | **RR within variant** | **Omicron, RR (95% CI)** |  | **Delta, RR (95% CI)** |  |
|  | Primary case vacc. status (16+) |  |  |  |  |
|  | Unvaccinated | Ref |  | Ref |  |
|  | Partial | 1.01 (0.64 - 1.58) |  | 0.99 (0.51 - 1.64) |  |
|  | Full | 1.04 (0.79 - 1.49) |  | 0.63 (0.46 - 0.89) |  |
|  | Boosted | 0.99 (0.68 - 1.49) |  | 0.18 (0.01 - 0.70) |  |
|  | Age of contacts |  |  |  |  |
|  | <16 years | 1.28 (0.97 - 1.83) |  | 1.35 (0.83 - 2.60) |  |
|  | 16-44 years | 1.16 (0.87 - 1.65) |  | 1.05 (0.65 - 2.03) |  |
|  | 45-64 years | 0.97 (0.73 - 1.39) |  | 1.14 (0.69 - 2.23) |  |
|  | 65+ years | Ref |  | Ref |  |
|  | Gender of contacts (16+) |  |  |  |  |
|  | Female | Ref |  | Ref |  |
|  | Male | 0.90 (0.79 - 1.03) |  | 1.03 (0.82 - 1.29) |  |
|  | Time since last vacc., (16+)<br>full vacc. contacts |  |  |  |  |
|  | ≤ 6 months | 0.89 (0.60-1.19) |  | 0.65 (0.34-1.07) |  |
|  | >6 months | Ref |  | Ref |  |

VE for booster vaccinated adult contacts was lower for Omicron (45%; CI_95_: 26-57) compared to Delta (65%; CI_95_: 42-80) but higher than for fully vaccinated (Table 1B). In the latter group, we found the protection against infection with Omicron to be 27% (CI_95_: 6-49), whereas 42% (CI_95_: 23-55) for Delta. Fully vaccinated primary cases have the same risk as unvaccinated primary cases in Omicron transmission to their adult household members (RR: 1.04; CI_95_: 0.79 - 1.49), Table 1C. The same pattern is observed for the booster vaccinated versus unvaccinated primary cases (RR: 0.99; CI_95_: 0.68 - 1.49). In contrast, booster vaccinated primary cases with Delta have 80% lower risk of Delta transmission (RR: 0.18; CI_95_: 0.01 - 0.70) relative to the unvaccinated primary cases.

When estimating the risk of infection, stratified by age groups, gender and the time since last dose (in fully vaccinated contacts), we found no significant differences (Table 1C).

## DISCUSSION

During a period when both the Omicron and Delta were circulating, we found an overall higher household ten-day secondary attack rate (SAR) for the Omicron (51%) compared to the Delta (36%) variant. This finding aligns with observations from Denmark [7] and the UK [14]. The SAR estimates were generally higher in our study, which could be due to various reasons such as differences in the testing regimes or discrepancies in the capacity and procedures for registering household contacts; see Supplementary S4 for study limitations. In this study, we have used contact tracing data, which may give higher estimates of SAR than registry-based studies since the exposure is verified through personal interviews. Furthermore, since the included children <16 years mainly were unvaccinated or partially vaccinated, this could contribute to the overall higher household SAR for both Delta and Omicron observed in our study. The overall SAR was substantially higher in households infected with Omicron, in line with the rapid takeover of Omicron from Delta in Norway. However, household exposure is often prolonged and repeated compared to social contacts in society, and preferably a complete evaluation should consider all close contacts. Unfortunately, the inclusion of non-household close contacts was not possible due to variations in contact tracing practices during the study period and between localities.

Unlike Lyngse et al., our study shows a significantly higher SAR for Omicron than Delta among unvaccinated household members, suggesting that intrinsic transmissibility is higher for the Omicron variant. This finding supports early assumptions that the Omicron is fundamentally more transmissible than Delta [15]. We also found a higher household SAR for Omicron among fully vaccinated and booster vaccinated household contacts when compared to Delta, indicating that immune evasion contributes to the increased transmissibility of the Omicron variant. The group of partially vaccinated individuals is heterogeneous and with varying vaccination dates, which may explain the lower RR of Omicron infection versus Delta compared to the other vaccination groups. Also, adolescents aged 12-19 years were overrepresented in this group. However, because we did not adjust for age in this analysis, our results should be interpreted cautiously.

Vaccination with two and three doses seemed to give lower protection against infection for Omicron than Delta, which is supported by other epidemiological studies and neutralization studies, likely related to a large number of mutations in the spike (S) protein compared to Delta [9]. While the protective effect of booster dose against Omicron infection (VE) was significant, the protection against onwards transmission seems rather low, compared to Delta. Our results indicate that booster vaccinated primary cases had approximately a four-fold higher risk of transmitting Omicron to their household contacts relative to Delta, which is higher than the risk of Omicron transmission versus Delta in fully vaccinated and unvaccinated primary cases. However, we did not adjust for age and time since vaccination, and thus, our results should be interpreted with caution.

In conclusion, our study indicates that the higher overall SAR among household contacts of Omicron cases is most likely due to higher intrinsic transmissibility of this variant and lower vaccine effectiveness. As reported by others, booster doses decrease this risk of infection with Delta and Omicron, but our findings suggest that it has limited effect on preventing Omicron transmission.

## Data Availability

The complete data set referred to in this manuscript is not publicly available as it contains person sensitive information. The Norwegian Institute of Public Health is data controller for the emergency preparedness registry BERECT-C19. The registry is temporary, and there are strict access control and routines for information handling information in the registry. Individual requests for access to non-sensitive data can be made to the Norwegian Institute of Public Health.

## Notes

## Acknowledgements

We are grateful to the Norwegian municipalities and their local contact tracing teams for gathering and sharing of data, and to the Norwegian emergency preparedness registry Beredt C19 for access to data.

## Disclaimer

No funding bodies had any role in study design, data collection and analysis, decision to publish, or preparation of the manuscript.

