## Supplementary Information (SI) for "Increased household transmission and immune escape of the SARS-CoV-2 Omicron variant compared to the Delta variant: evidence from Norwegian contact tracing and vaccination data"

### Supplementary S1: Data sources

We used data from the Norwegian COVID-19 pandemic preparedness registry, Beredt C19 containing individual level information from Norwegian health registries. The following Beredt C19 data sources were used in this study:

- Norwegian vaccination registry (SYSVAK)
  - Vaccination date
  - Vaccine type
- Norwegian Surveillance System for Communicable Diseases (MSIS)
  - Test date
  - Positive test result for SARS-CoV-2, either by RT-PCR or antigen rapid test.
  - Virus variant, detected through whole genome sequencing (WGS) or PCR variant screening.
- Norwegian Population Registry
  - Age
  - Birth
  - Gender
- Municipal contact tracing data (KS Fiks Smittesporing) from 64 Norwegian municipalities in 11 different counties. Only data from 57 municipalities fulfilled the inclusion criteria and were included in the study.
  - Linkage between index cases and their corresponding close contacts, identified through manual contact tracing by trained personnel in the municipalities.
- Statistics Norway (SSB)
  - Household ID key

### Supplementary S2: Definitions

Household primary case: The first person in the household who has tested positive for SARS-CoV-2 in the period from 14 December 2021-13 January 2022 and was registered as an index in the contact tracing data.

Household member: All individuals registered in the same household, based on Statistics Norway (SSB) household data.

Household contact: Contacts identified by the contact tracing team, registered in the contact tracing system, and matched with Statistics Norway (SSB) to determine that they belong to the same household as the primary case.

Registration date: Date of registration in the contact tracing system.

Household secondary case: An individual registered as a close contact of the primary case, living in the same household and who had test date within and including ten days after the test date of the primary case.

Household secondary attack rate (SAR): The probability that an infectious person infects a person in the same household during his or her infectious period. We considered the infectious period of the primary cases as ten days from their positive test dates, meaning we calculated the SAR10 in our analysis.

Variant identification: The variant was determined by the PCR screening, whole genome sequencing (WGS), or both methods.

To define the vaccine status of the household contacts, we used the test date of the primary case and compared it with the contacts vaccination dates:

1. Unvaccinated: A contact was considered unvaccinated if the primary case's test date is before the contact's first dose.
2. Partially vaccinated: A contact was considered partially vaccinated if he/she had received 1 dose of vaccine (mRNA Vaccines or AstraZeneca vaccine) prior to the test date of his/her primary case. Contacts who had received dose 2 within the last week before the primary case's test date were also considered partly vaccinated.
3. Fully vaccinated: A contact was considered fully vaccinated if he/she had received dose 2 (mRNA) at least 1 week prior to the test date of his/her primary case.
4. Booster vaccinated: A contact was considered booster vaccinated if he/she had received dose 3 at least 1 week prior to the test date of his/her primary case. The time interval between the second and the third doses should be >= 120 days.

The vaccine status of the primary cases was defined based on their test date and their vaccination dates. Individuals with J&J vaccine were excluded from the study. We excluded households where two individuals tested positive on the same day to ensure a unique index case in each household.

Supplementary S3: Study sample characteristics by age and vaccination status

| **Table S3A Omicron Households: Age and vaccination status of primary cases and contacts** (n=667 primary cases, n= 1299 contacts). | | | | | | | | | | | | | | | | |
|---|---|---|---|---|---|---|---|---|---|---|---|---|---|---|---|---|
| | Vaccination status primary cases | | | | | | | | Vaccination status contacts | | | | | | | |
| | Unvaccinated | | Partially vaccinated | | Fully vaccinated | | Booster vaccinated | | Unvaccinated | | Partially vaccinated | | Fully vaccinated | | Booster vaccinated | |
| Total | 151 | | 69 | | 372 | | 75 | | 322 | | 121 | | 654 | | 202 | |
| Age groups | n | % | n | % | n | % | n | % | n | % | n | % | n | % | n | % |
| 0-15 | 109 | 0.712 | 43 | 0.281 | 1 | NA | NA | NA | 278 | 0.781 | 78 | 0.219 | NA | NA | NA | NA |
| 16+ | 42 | 0.082 | 26 | 0.051 | 371 | 0.721 | 75 | 0.146 | 44 | 0.047 | 43 | 0.046 | 654 | 0.692 | 202 | 0.214 |

| **Table S3B. Delta households: Age and vaccination status of primary cases and contacts** (n=455 primary cases, n= 870 contacts). | | | | | | | | | | | | | | | | |
|---|---|---|---|---|---|---|---|---|---|---|---|---|---|---|---|---|
| | Vaccination status primary cases | | | | | | | | Vaccination status contacts | | | | | | | |
| | Unvaccinated | | Partially vaccinated | | Fully vaccinated | | Booster vaccinated | | Unvaccinated | | Partially vaccinated | | Fully vaccinated | | Booster vaccinated | |
| Total | 202 | | 35 | | 204 | | 14 | | 297 | | 62 | | 445 | | 66 | |
| Age groups | n | % | n | % | n | % | n | % | n | % | n | % | n | % | n | % |
| 0-15 | 141 | 0.876 | 20 | 0.124 | NA | NA | NA | NA | 236 | 0.822 | 49 | 0.171 | 2 | NA | NA | NA |
| 16+ | 61 | 0.207 | 15 | 0.051 | 204 | 0.694 | 14 | 0.048 | 61 | 0.105 | 13 | 0.022 | 443 | 0.760 | 66 | 0.113 |

**Table S3C:** Number of all contacts (N), and infected contacts (n) of Omicron versus Delta stratified by contacts' age, primary cases' age, vaccination status of contacts, primary cases, gender, and time since last vaccination in fully vaccinated contacts in Norway, 14 December 2021 to 23 January 2022.

| | Omicron | | | Delta | | |
|---|---|---|---|---|---|---|
| Description | All contacts (N) | Infected contacts (n) | Percent (n*100/N) | All contacts (N) | Infected contacts (n) | Percent (n*100/N) |
| Overall | 1299 | 662 | 50.89 | 870 | 315 | 36.21 |
| Age of contacts | | | | | | |
| <16 years | 356 | 206 | 57.87 | 287 | 120 | 41.81 |
| 16+ years | 943 | 455 | 48.25 | 583 | 195 | 33.45 |
| Age of primary cases | | | | | | |
| <16 years | 354 | 188 | 53.11 | 353 | 123 | 34.84 |
| 16+ years | 945 | 473 | 50.05 | 517 | 192 | 37.14 |
| Vacc. status, contacts | | | | | | |
| Unvaccinated | 322 | 190 | 59.01 | 297 | 138 | 46.46 |
| Partial | 121 | 69 | 57.02 | 62 | 21 | 33.87 |
| Full | 654 | 326 | 49.85 | 445 | 143 | 32.13 |
| Boosted | 202 | 76 | 37.62 | 66 | 13 | 19.70 |
| Vacc. status, primary cases | | | | | | |
| Unvaccinated | 329 | 187 | 56.84 | 420 | 158 | 37.62 |
| Partial | 154 | 64 | 41.56 | 71 | 28 | 39.44 |
| Full | 713 | 363 | 50.91 | 360 | 127 | 35.28 |
| Boosted | 103 | 47 | 45.63 | 19 | 2 | 10.53 |
| Contact vacc. status (16+) | | | | | | |
| Unvaccinated | 44 | 30 | 68.18 | 61 | 34 | 55.74 |
| Partial | 43 | 23 | 53.49 | 13 | 5 | 38.46 |
| Full | 654 | 326 | 49.85 | 443 | 143 | 32.28 |
| Boosted | 202 | 76 | 37.62 | 66 | 13 | 19.70 |
| Primary case vacc. status (16+) | | | | | | |
| Unvaccinated | 51 | 23 | 45.10 | 61 | 29 | 47.54 |
| Partial | 44 | 20 | 45.45 | 17 | 8 | 47.06 |
| Full | 499 | 235 | 47.09 | 235 | 70 | 29.79 |
| Boosted | 83 | 37 | 44.58 | 12 | 1 | 8.33 |
| Age of contacts | | | | | | |
| <16 years | 356 | 206 | 57.87 | 287 | 120 | 41.81 |
| 16-44 years | 497 | 259 | 52.11 | 365 | 119 | 32.6 |
| 45-64 years | 395 | 173 | 43.8 | 189 | 67 | 35.45 |
| 65+ years | 51 | 23 | 45.1 | 29 | 9 | 31.03 |
| Gender of contacts (16+) | | | | | | |
| Female | 513 | 259 | 50.49 | 294 | 97 | 32.99 |
| Male | 430 | 196 | 45.58 | 289 | 98 | 33.91 |
| Time since last vacc., (16+) full vacc. contacts | | | | | | |
| ≤ 6 months | 45 | 20 | 44.44 | 46 | 10 | 21.74 |
| >6 months | 609 | 306 | 50.25 | 397 | 133 | 33.50 |

### Supplementary S4: Data Limitations

Municipalities have used different approaches to contact tracing and testing of contacts based on local priorities, resources and regulations. This means that the quality of the contact tracing data will vary between municipalities and over time. We have therefore restricted our analyses to household contacts, as the contact tracing teams have generally prioritized them. However, we cannot rule out that even some household contacts have not been registered in the contact tracing system, particularly in periods of very high incidence. Municipalities were recommended to prioritize contact tracing around cases with a high risk of severe illness or high risk of transmission to many contacts in situations with particularly high incidence.
Since exposure within the same household is often prolonged over several days, the estimated secondary attack rate might reflect several transmission chains. Also, since we cannot distinguish asymptomatic from symptomatic infection in our data set, we are not able to assess the protective effect of vaccination against disease.
We assume that most household contacts were tested at least once during their ten days of quarantine, especially since a negative test result on day seven would allow termination of quarantine. However, since we did not include the test record of the contacts (including any positive result of antigen-self tests not confirmed in the national surveillance system), we cannot rule out that the true number of infected contacts might be higher than captured in this study.

In contrast to pure registry-based data, contact tracing data, provides qualitative data on true exposures identified through manual contact tracing (interviews). Ideally, contact tracing data should be combined with either "contact category" (e.g. household contact, friend, colleague etc.), or updated information on place of living, to separate household contacts from "other contacts". We matched contact tracing data with household registry information to identify which contacts belonged to the same household. Since we did not have access to either contact category or current living address, the actual number of household contacts might diverge from our estimates.
Regarding our assessments on the effect of vaccination status and waning, we have not included detailed information on the time since last vaccination or vaccine types administrated. These characteristics should be more thoroughly investigated in separate studies focusing on VE.
